# Unraveling the role of rise in temperature on the emergence of antimicrobial resistance

**DOI:** 10.1101/2023.09.06.23295147

**Authors:** Anuradha Goswami, J. Jeffrey Morris

**Author notes:** Alternate correspondence. Address: 3100 East Science Hall, 902 14th Street South Department of Biology, University of Alabama at Birmingham, Birmingham, AL 35294-1170.

## Abstract

The ability of bacteria to resist the effects of antibiotics, or antimicrobial resistance (AMR), is a growing risk to world health, making it more challenging to combat infectious health problems. The growth rate of bacteria is significantly influenced by temperature, particularly temperatures between 35 and 37°C, often considered the most suitable for the spread of human illnesses. Knowing that a rise in temperature influences bacterial growth rate, contributing to a higher infection rate, it is imperative to unravel and comprehend the association between climate change and the emergence of AMR. We hypothesized that rising temperatures could exacerbate the emergence of AMR in opportunistic and pathogenic bacteria. To test our hypothesis, we investigated the global distribution of AMR and the correlation between AMR and socioeconomic factors, climate change, and air quality in the United States. The study found high resistance rates to common infections such as Methicillin-resistant Staphylococcus aureus (MRSA) and Vancomycin-resistant enterococcus (VRE) infections are prevalent in many countries. In the United States, MRSA-AMR was more common in low-income states with increased poverty rates and poor air quality. The study also found a positive correlation between the rise in temperature over the past 10 years and AMR bacterial infections. The investigation concluded that socioeconomic factors, climate change, and race collectively impact the prevalence of AMR infections. The probability of AMR infection upsurging in the next decade was highest within states with more frequent rises in temperature over the last 10 years. The model predicted that states with at least 1 °C rise in temperature over the previous 10 years are expected to experience a surge in AMR bacterial infections in coming years. Though the statistical details might vary depending on the data collected in future, the correlation between climate change and the emergence of AMR in bacterial infection is alarming. The study indicates that climate change has an essential, largely unrecognized influence on AMR bacterial infections that warrants additional research. It implies that comprehensive and integrated strategies are needed to address the AMR and climate change challenges.

## 1. INTRODUCTION

Global health is significantly at risk from climate change. Extreme weather, sea level rise, changes in agricultural yields, ecosystem damage, declining microbial biodiversity, and increased disease transmission are some of the most significant threats climate change brings [1]. The increased emergence rate of antimicrobial resistance (AMR) genes in the environment is one of the most underappreciated threats of climate change [2]. Elevated temperatures may change the genetic diversity of microbial communities as certain species adapt more rapidly to the changing climate than others, and AMR may increase simultaneously in several ways. For instance, rising temperatures enable microorganisms formerly restricted to tropical regions to thrive in temperate zones, potentially resulting in the appearance of new AMR bacteria and the spread of AMR into native bacteria via horizontal gene transfer from new colonizing taxa carrying exotic AMR genes. Climate change might also induce stress in bacteria, which increases their risk of developing AMR. For instance, a drier environment might cause desiccation stress in microorganisms, selecting for resistance phenotypes that may also confer AMR and other pathogenesis phenotypes [3-5]. AMR infectious agents prevalent in patients across the world and included in this study are:

### Methicillin-resistant *Staphylococcus aureus* (MRSA)

is a complex of widespread *S. aureus* strains defined by their broad resistance to the standard antibiotics used to treat typical staph infections, leading to the development of recalcitrant infections [6]. Most MRSA infections affect patients who have visited hospitals or other healthcare facilities, like nursing homes and dialysis facilities; when encountered in such environments, it is referred to as healthcare-associated MRSA (HA-MRSA). Invasive operations or equipment, such as surgeries, intravenous tubing, or prosthetic joints, are frequently linked to HA-MRSA infections. Another kind of MRSA infection has been reported among healthy individuals in the larger community. Community-associated MRSA (CA-MRSA) often starts as an uncomfortable skin boil that can diffuse in a community by skin-to-skin contact. People who live in crowded housing and childcare workers are examples of at-risk populations [7].

### Vancomycin resistance *Enterococcus* (VRE)

has developed resistance to the “last resort” intravenous drug Vancomycin. Like MRSA, VRE infections often occur in healthcare facilities such as hospitals and have been linked to institutional outbreaks. Also, like MRSA, the bacteria frequently colonize people as commensals without producing symptoms [8]. To stop transmission, some hospitals segregate patients who carry commensal VRE [9, 10].

### Third-generation cephalosporin-resistance (3GCR)

Cephalosporins are a class of synthetic molecules containing the bactericidal beta-lactam structure found in natural penicillin. Because of their altered molecular shape, these drugs can counter infections resistant to penicillin and other common antibiotics. The synthesis of extended-spectrum beta-lactamase (ESBL) is frequently the cause of resistance to the most recent crop of third-generation cephalosporins, and bacteria that can produce these enzymes are becoming more common in *Escherichia coli* and *Klebsiella pneumoniae*, and other opportunistic pathogen species [11, 12]. ESBLs are active against all beta-lactam drugs, such as penicillin and cephalosporins, rendering them useless for treating 3GCR infections [13]. 3GCR organisms are a rising cause of sickness in hospitals and the general population and are frequently treated using non-beta-lactam antibiotics. Similar to VRE, these microorganisms can colonize the gastrointestinal tract asymptomatically and produce symptomatic disease when the opportunity arises [8].

### Carbapenem-resistant Enterobacteriaceae (CRE)

have genetic mechanisms of resistance that confer resistance to practically all beta-lactams, including the broad-spectrum class of carbapenems [14]. Despite being widespread, several mechanisms of carbapenem resistance are spreading quickly from areas with high antimicrobial usage and resistance, such as OXA-48 and New Delhi metallo-beta-lactamase-1 (NDM-1) beta-lactamases [15, 16]. Like the strains described above, CRE can cause clinical disease and asymptomatic gut colonization and antibiotics other than beta-lactams are frequently utilized to treat these pathogens.

The summer months experience an upsurge in bacterial infections worldwide [17]. It is well-recognized that temperature is essential in regulating microbial activity and affecting microbial populations [18]. However, research is still needed to determine whether AMR in opportunistic pathogenic bacteria and climate change are interrelated. In this study, we perform a meta-analytic evaluation of AMR concerning socioeconomic and climatic change issues. We hypothesize that increases in average temperature positively correlate to the rate of AMR in pathogenic bacteria. We specifically pursue the following objectives:

1. To determine the global distribution of MRSA, 3GCR, VRE, and CRE.
2. To investigate the relationship between AMR, climate change (i.e., rise in average temperature between 2013 and 2022), and socioeconomic factors in the United States and
3. To predict the states in the United States most vulnerable to AMR infections and rise in temperature in coming years.

## 2. METHODS

### 2.1. Worldwide patient bacterial isolate AMR phenotype data

Publicly available data was used to investigate the current level of AMR in connection with socioeconomic and climate change factors. We used the HealthMap ResistanceOpen database [19] to assemble information on the prevalence of MRSA, 3GCR, VRE, and CRE bacterial isolates collected from outpatient and inpatient infection samples from 2013 to 2022 in the United States, The United Kingdom (UK), Brazil, Canada, and European Union nations (EU). The percentage of patient isolates with an AMR phenotype, and the total number of isolates used in the analysis are given in the supplemental information. The United States was used as the study’s model for further examination to test the research premise.

### 2.2. State-wide rise in temperature in the United States from 2013 to 2022

The National Centers for Environmental Information National Oceanic and Atmospheric Administration was used to obtain the average temperature for each month in 2013 and 2022 across the United States [20]. The state-wide rise in temperature in the United States in the last ten years was calculated by finding the difference between each month’s average temperature in 2013 and 2022. The detailed table of temperature rise values used for the analysis is given in supplemental information.

### 2.3. Air quality state-wide ranking and other socioeconomic factors in the United States

Air Quality by State, a publicly available database provided by Wisevoter [21], was used to find the state-wide air quality data. The states were ranked based on the percentage of unhealthy air quality days from 1 to 52, where rank 1 corresponds to 0% unhealthy air quality. The socioeconomic factors across the United States include Poverty percentage (2021), Median household income in 2015, percentage of black or African American population, state-wide population for age above 65 years and obesity percentage taken from US Census Quick Facts [22].

### 2.4. Geospatial mapping of AMR bacterial infection

To create a geospatial map illustrating the prevalence of AMR infection, the percentage of bacterial cultures that contained MRSA, 3GCR, VRE, and CRE in the United States, European Union nations, and other nations (Brazil, Canada, and UK) was used as an input variable. The thematic map data, including details on country borders, coordinates, and other geographic aspects and observed AMR data, were combined using the region variable (a common variable that appears in both data sets). The thematic map data was used to create the base map, which showed the borders of countries and regions. The observed AMR data created the layers overlaid on the base map. The color of each layer displayed the proportion of AMR bacterial cultures. The R codes used for the geospatial mapping of AMR prevalence are mentioned in the Supplementary files.

### 2.5. Prediction and evaluation of the rate of AMR infection across the United States

A generalized linear model *glm()*, was fitted to predict the rate of occurrence of AMR bacterial infection (dependent variable) based on the independent variable-(*i*) average rise in temperature (from 2013 to 2022), (*ii*) number of bacterial isolates cultured from patients across the United States and (*iii*) geographical location (regions) of the states. The model assumes that the average AMR infection follows a Gaussian distribution and that the logarithm of the mean of the average AMR infection is a linear function of the independent variables. The logarithm of the mean of the dependent variable was evaluated as a linear function of the independent variables where the effect and interaction of each independent variable were fitted in the model. A data frame was created to anticipate the rate of introduction of AMR infections over the following ten years based on the model coefficient and the existing trend of temperature rise. The model was trained on the new data frame constructed and then was used to predict an average number of AMR bacterial infections across the states in the United States. The R codes used for model construction, training and prediction are mentioned in the Supplementary files. The median observed (O_AMR_)and predicted (P_AMR_)percentage of the average number of bacterial isolates with AMR (Average of MRSA, 3GCR, VRE and CRE %) was used to calculate the increase in the rate of occurrence of AMR infection (*r*_*n*_) per year for n years (n = 10)

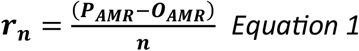

The rate of AMR infections in patients expected to occur in each state in the United States during the next ten years was calculated as follows.

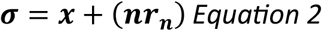

*Where x is the current value of the proportion of bacterial isolates with AMR phenotype in each state of the United States*.

The two-layer data filtering was applied to examine the direct relationship between a rise in temperature and an AMR bacterial infection. States having an average temperature difference (or rise in temperature) between 2013 and 2022 that was larger than or equal to 0.5°C were first filtered. From the filtered data, states with an average proportion of AMR bacterial infections greater than or equal to the median value (=14.75%) were then classified. The final data frame was utilized as an input variable to assess the effect of the rate of temperature change over time on the prevalence of AMR infection in the United States. By fitting a linear model utilizing the average temperature of each state over 10 years from 2013 to 2022 as the dependent variable, the slope (=rate of change in temperature of states over time) was evaluated in R. Regression plots were made using the *ggplotRegression()* function from the *ggplot2* library. Then, as described above, a generalized linear model was built using the slope of each state’s average temperature time series model as a variable.

## 3. RESULTS

### 3.1. Spatial mapping of AMR bacterial infections in global patient samples

The global distribution of MRSA, 3GCR, VRE, and CRE for the United States and European nations and the overall distribution are each shown in Figure 1(a-c), respectively. The spatial mapping revealed that a sizeable proportion of *S. aureus* isolated from inpatient and outpatient samples worldwide were methicillin resistant. The 3GCR *E. coli* and CRE *K. pneumoniae* percentag*e* was higher in the EU, UK, and Brazil. *S. aureus* infections with MRSA isolates have become prevalent throughout the US and Brazil in the past ten years. Nonetheless, it was considerably less widespread in the European nations. 3GCR in Europe and other countries studied were similar except for Greece, where a relatively higher proportion of 3CGR infections were mapped. However, VRE and CRE were more prevalent in EU countries, especially France (both), Poland (VRE), Romania, Bulgaria, and Greece.

**Figure 1.**
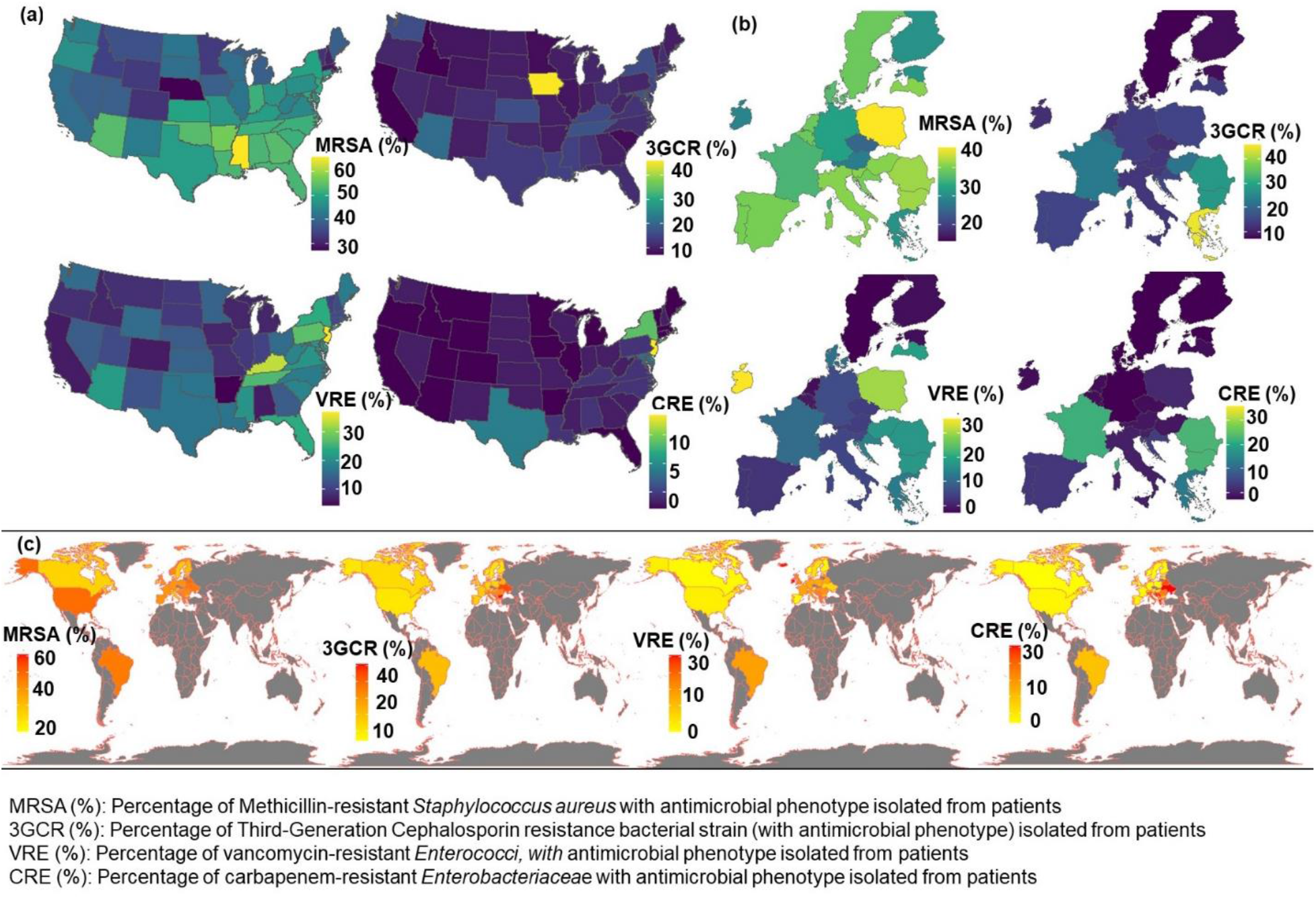
Geospatial mapping of the percentage of MRSA, 3GCR, VRE, CRE resistant bacterial isolates (a) United States of America ^§^; (b) Only Europe; (c) Worldwide. ^§^: Alaska used in the analysis is not displayed in the United States heatmap.

The United States was determined to have the highest prevalence of MRSA infections. Inpatient and outpatient samples of S. aureus from Alabama, Arizona, Arkansas, Connecticut, Florida, Georgia, Indiana, Kansas, Kentucky, Louisiana, Maryland, Mississippi, New Jersey, New York, North Carolina, Oklahoma, South Carolina, Tennessee, and Texas in the United States revealed a significant percentage (45% and above) of methicillin-resistant (MRSA) strains. However, at least 10% of the bacterial isolates in the patient samples from Arizona, Florida, Georgia, Kansas, Kentucky, Louisiana, Maryland, Mississippi, and Tennessee were resistant to vancomycin.

### Correlation between socioeconomic variables, air quality, AMR and temperature rise in the United States

We hypothesized that the rate of AMR will differ according to economic situation, population demography (age and race), pollution level (air quality), and/or rise in temperature. To separate the relationships between factors linked to the rise in MRSA infections in patients across the United States, the socioeconomic and environmental variables were split into three groups, namely, (*i*) state-wide economic status, (*ii*) state-wide population and (*iii*) environmental influence and other factors. The MRSA was chosen since prior geospatial mapping suggested it was more common in the United States than other AMR-phenotypes in patient bacterial cultures. The percentage of bacterial isolates with AMR phenotype was significantly positively correlated with states with low income and high poverty (Figure 2 (a) Supplementary Figure 1). Moreover, states with low air quality were significantly positively correlated with MRSA infection across the states (Figure 2 (a), Supplementary Figure 1). The study population included out and inpatients’ data with MRSA infections collected in the United States between 2013 and 2022. The states with the highest prevalence of MRSA infection were Mississippi, Arkansas, North Carolina, Nebraska, Kansas, Kentucky, and Tennessee, which also had an average of 0.93±0.65°C rise in temperature (June-to-September) since 2013 compared to a nationwide average of 1.30±0.81°C. The study also found that the proportion of a state’s Black or African American population significantly correlated with an increased rate of MRSA infections (Figure 2 (a-b), Supplementary Figure 1). Thus, it can be suggested that Black or African American patients are more at risk of being infected with MRSA than the white population (Supplementary Figure 1) in the United States.

**Figure 2.**
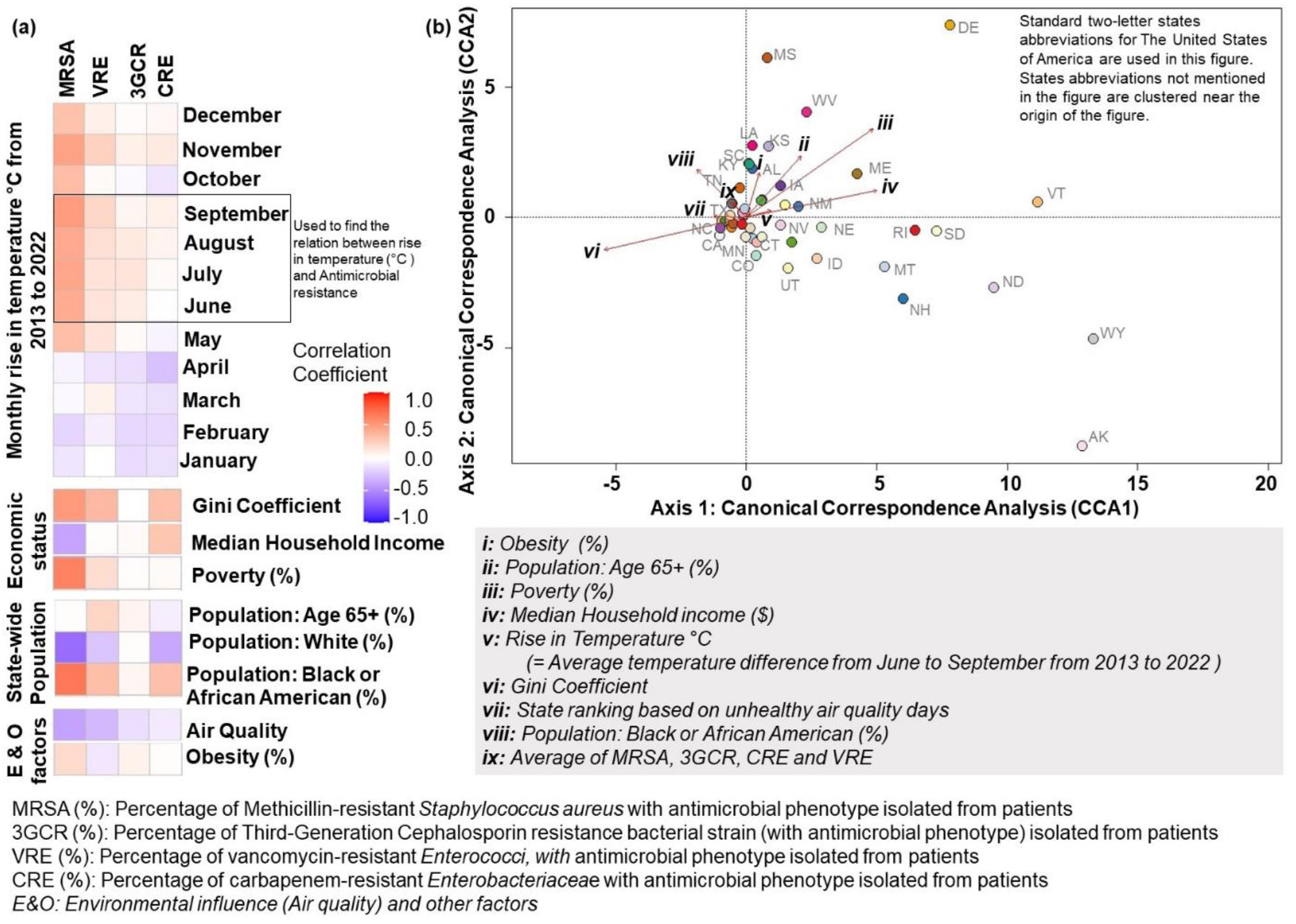
**(a)** Correlation between percentage of bacterial isolates from patients exhibiting antimicrobial resistance (AMR) phenotype and time series state-wide rise in temperature in the United States; **(b)** Multivariate statistical analysis depicting the association between AMR and state-wide physical-socioeconomic diversity in the United States. Note: Supplementary Figure (Figure 1) illustrates linear regression of the fitted model for state-wide economic status, population, air quality and obesity percentage across the United States, respectively.

The data revealed that an extensive number of bacterial infections with the AMR phenotype arose during the summertime (Figure 2(a)); therefore, a state-wide rise in temperature in a decade (difference between temperature in 2022 and 2013) from June to September was used to evaluate the study’s notion. The multivariate statistical analysis explored the variables (shown as arrows in Figure 2(b)) contributing to the state-wise emergence of AMR bacterial infections across the United States in the last ten years. With a decline in environmental quality and temperature rise, residents of states with high poverty rates and low household incomes are more likely to become infected with AMR. These findings support the study’s hypothesis that there is a connection between poverty, global warming, and AMR. Due to climate change, low-income states are more likely to endure rising temperatures and have less access to healthcare. Controlling the spread of MRSA and other AMR bacteria may become more challenging due to the interaction of these factors. The results of this study suggest that socioeconomic factors, climate change, and AMR in the United States are intricately linked. These findings need further investigation and discussion, considering they have significant health implications for the general population.

### 3.3. Analyzing effects of temperature rise on the emergence of antibiotic resistance

#### (a) AMR in the United States stratified by state and rise in temperature

To train the GLM model to predict the effects and interactions of the average temperature increase between 2013 and 2022, the number of bacterial isolates tested, the areas or locations of the states (depending on its direction), and the rate of AMR infection, average monthly (June to September) temperature difference (2013 and 2022) data was used. The influence of temperature increase and the quantity of state-specific bacterial isolates were favorably linked (p-value = 0.6) with the percentage of AMR bacterial infections examined. To predict the rate of AMR infection assuming a comparable trend of temperature rise, the data frame of response variables was generated as indicated in Methods 2.5. The model’s fitted values were tightly spaced between 2.5 and 3.0, indicating the relative accuracy of the model’s prediction (Supplementary Figure 2).

Whereas the observed (present) median value of AMR bacterial infection was 14.8%, the model predicted that after ten years, a median increase of 17.5% could occur (Figure 3(a)). Assuming a constant rate of change in temperature and other variables, Equation 1 predicts a yearly increase in AMR infections of 0.27%. The state-specific observed and anticipated rise in AMR infection is illustrated in Figure 3(b). According to the model, 28 of the 49 states studied are at risk of witnessing an increase in the prevalence of AMR infections during the next ten years (Table 1). The expected AMR infections (average (MRSA, 3GCR, CRE, and VRE) %) because of the increase in temperature in the United States are presented in Figure 3 (c). The scatter plot demonstrates a statistically significant difference in the expected AMR infection rate between states with a temperature rise of less than 1°C and more than or equal to 1°C (*T-test p-value = 0*.*02*). Thus, it can be inferred that states experiencing frequent temperature variations are more likely to experience an upsurge in AMR infections in the future.

**Table 1.**
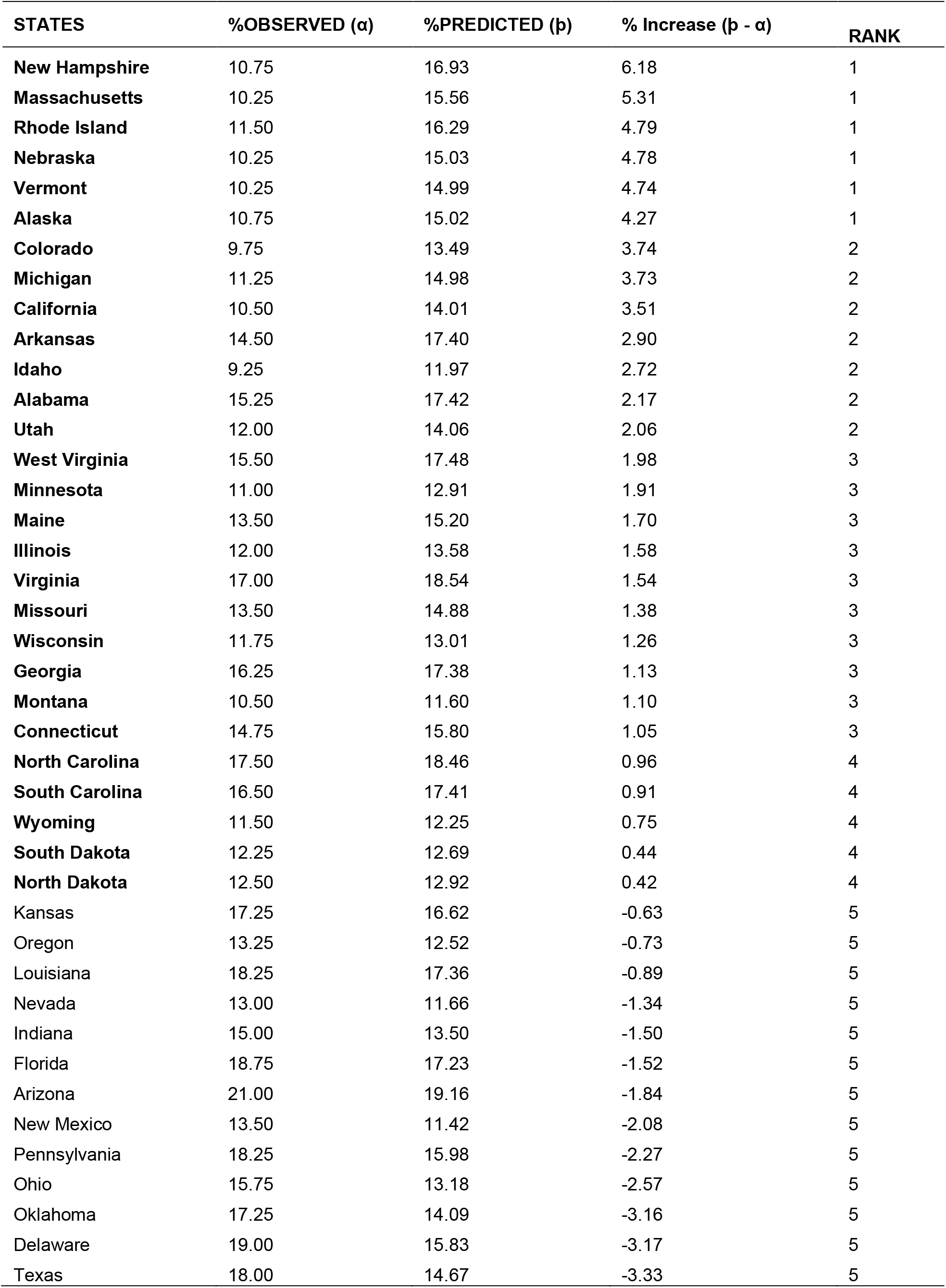
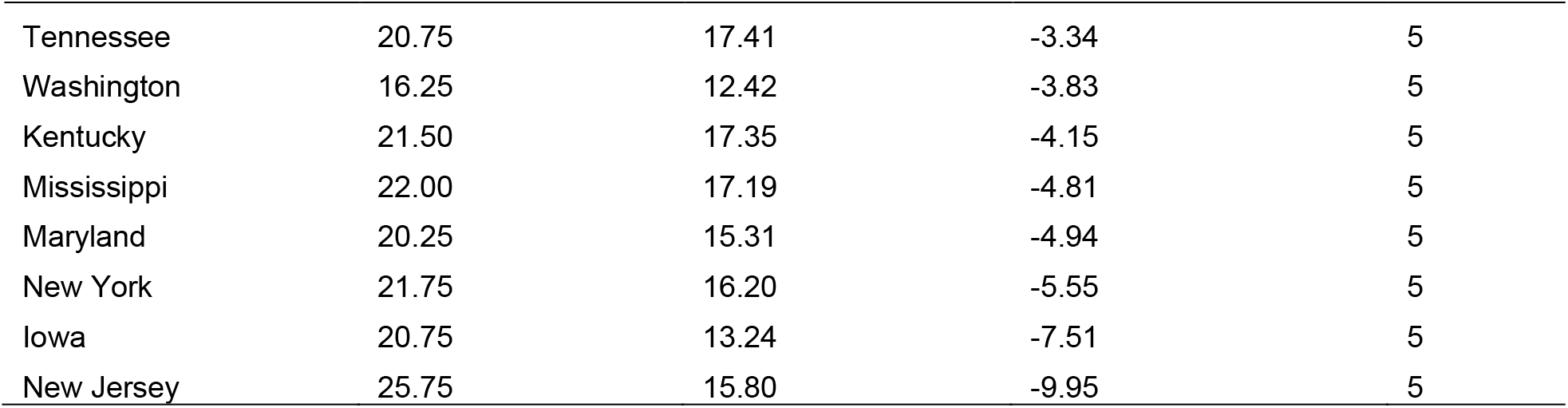
List of states with observed (current) and predicted AMR bacterial infection in ten years. The states in bold in the table are at high risk of increasing AMR infection rates. The states are ranked based on the severity of the risk: Rank 1 = high risk (> 4% predicted increase in AMR infection rate), Rank 5 = low risk. Note: The prediction data assumes that all factors used in the model will have a similar trend in a decade.

| STATES | %OBSERVED ($\alpha$) | %PREDICTED ($\beta$) | % Increase ($\beta - \alpha$) | RANK |
| --- | --- | --- | --- | --- |
| New Hampshire | 10.75 | 16.93 | 6.18 | 1 |
| Massachusetts | 10.25 | 15.56 | 5.31 | 1 |
| Rhode Island | 11.50 | 16.29 | 4.79 | 1 |
| Nebraska | 10.25 | 15.03 | 4.78 | 1 |
| Vermont | 10.25 | 14.99 | 4.74 | 1 |
| Alaska | 10.75 | 15.02 | 4.27 | 1 |
| Colorado | 9.75 | 13.49 | 3.74 | 2 |
| Michigan | 11.25 | 14.98 | 3.73 | 2 |
| California | 10.50 | 14.01 | 3.51 | 2 |
| Arkansas | 14.50 | 17.40 | 2.90 | 2 |
| Idaho | 9.25 | 11.97 | 2.72 | 2 |
| Alabama | 15.25 | 17.42 | 2.17 | 2 |
| Utah | 12.00 | 14.06 | 2.06 | 2 |
| West Virginia | 15.50 | 17.48 | 1.98 | 3 |
| Minnesota | 11.00 | 12.91 | 1.91 | 3 |
| Maine | 13.50 | 15.20 | 1.70 | 3 |
| Illinois | 12.00 | 13.58 | 1.58 | 3 |
| Virginia | 17.00 | 18.54 | 1.54 | 3 |
| Missouri | 13.50 | 14.88 | 1.38 | 3 |
| Wisconsin | 11.75 | 13.01 | 1.26 | 3 |
| Georgia | 16.25 | 17.38 | 1.13 | 3 |
| Montana | 10.50 | 11.60 | 1.10 | 3 |
| Connecticut | 14.75 | 15.80 | 1.05 | 3 |
| North Carolina | 17.50 | 18.46 | 0.96 | 4 |
| South Carolina | 16.50 | 17.41 | 0.91 | 4 |
| Wyoming | 11.50 | 12.25 | 0.75 | 4 |
| South Dakota | 12.25 | 12.69 | 0.44 | 4 |
| North Dakota | 12.50 | 12.92 | 0.42 | 4 |
| Kansas | 17.25 | 16.62 | -0.63 | 5 |
| Oregon | 13.25 | 12.52 | -0.73 | 5 |
| Louisiana | 18.25 | 17.36 | -0.89 | 5 |
| Nevada | 13.00 | 11.66 | -1.34 | 5 |
| Indiana | 15.00 | 13.50 | -1.50 | 5 |
| Florida | 18.75 | 17.23 | -1.52 | 5 |
| Arizona | 21.00 | 19.16 | -1.84 | 5 |
| New Mexico | 13.50 | 11.42 | -2.08 | 5 |
| Pennsylvania | 18.25 | 15.98 | -2.27 | 5 |
| Ohio | 15.75 | 13.18 | -2.57 | 5 |
| Oklahoma | 17.25 | 14.09 | -3.16 | 5 |
| Delaware | 19.00 | 15.83 | -3.17 | 5 |
| Texas | 18.00 | 14.67 | -3.33 | 5 |
| Tennessee | 20.75 | 17.41 | -3.34 | 5 |
| Washington | 16.25 | 12.42 | -3.83 | 5 |
| Kentucky | 21.50 | 17.35 | -4.15 | 5 |
| Mississippi | 22.00 | 17.19 | -4.81 | 5 |
| Maryland | 20.25 | 15.31 | -4.94 | 5 |
| New York | 21.75 | 16.20 | -5.55 | 5 |
| Iowa | 20.75 | 13.24 | -7.51 | 5 |
| New Jersey | 25.75 | 15.80 | -9.95 | 5 |

**Figure 3.**
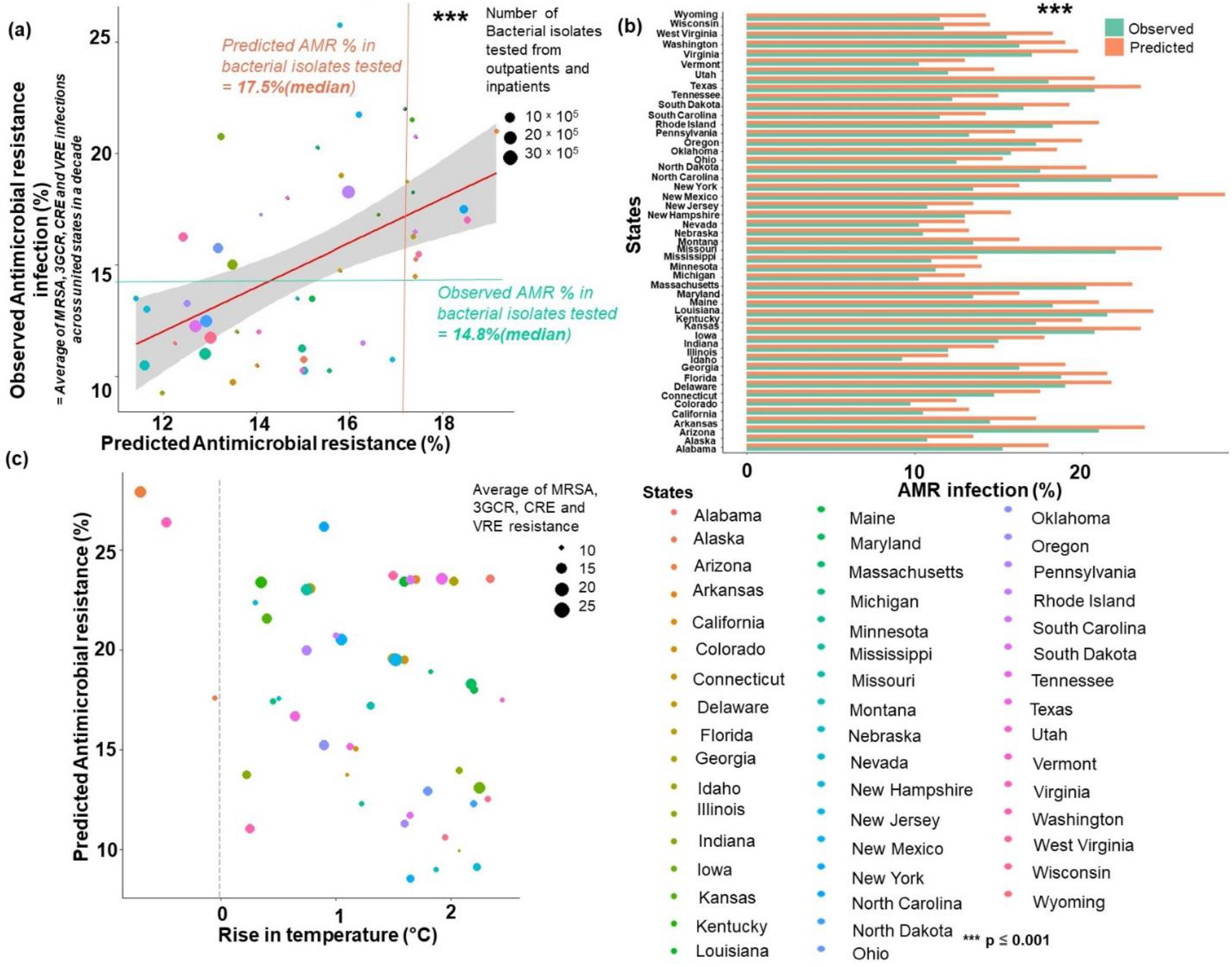
Predicted antimicrobial resistance (AMR) infection in the United States in a decade. **(a)** Linear regression of observed versus AMR bacterial infection in the United States; **(b)** Predicted AMR infection in a decade as a function of rise in temperature. **(c)** Predicted AMR infection in a decade, stratified by state. Note: The different colors represent the states of the United States.

### (b) Rate of change in temperature and AMR bacterial infections over time

The states that were a good fit to study the effects of the temperature rise and AMR bacterial infections were found using the two-layer data filtering and screening method described above in the methods section. The list of states that met the screening criteria and were further studied includes Alabama, Connecticut, Delaware, Florida, Georgia, Iowa, Louisiana, Maryland, Mississippi, New Jersey, New York, North Carolina, Ohio, Oklahoma, Pennsylvania, South Carolina, Tennessee, Texas, and West Virginia. The slope of a temperature time series linear model for each state over four-month intervals (June to September) throughout ten years from 2013 to 2022 (refer to Supplementary Figure 3) was used to measure the rate of change in AMR infection over time. The months were selected based on the previous experiment (Figure 2(a)), which showed that midsummer temperatures were ideal for the growth of the AMR bacterial infection. The GLM model (Figure 4) predicted a correlation between an upward trend in AMR infection and the rise in average temperatures in the US over ten years. Therefore, it could be justified to infer that an increase in temperature may promote bacterial growth and the development of AMR. Figure 4 demonstrates that the data are highly varied, and states tend to group at different axial positions (illustrated by shaded patches). These suggest that in addition to the increase in temperature, there could be additional factors, a few of which were previously emphasized, that contribute to the AMR infection. Therefore, the rise in AMR infection is a comprehensive effect of many variables and requires a holistic approach for further investigation.

**Figure 4.**
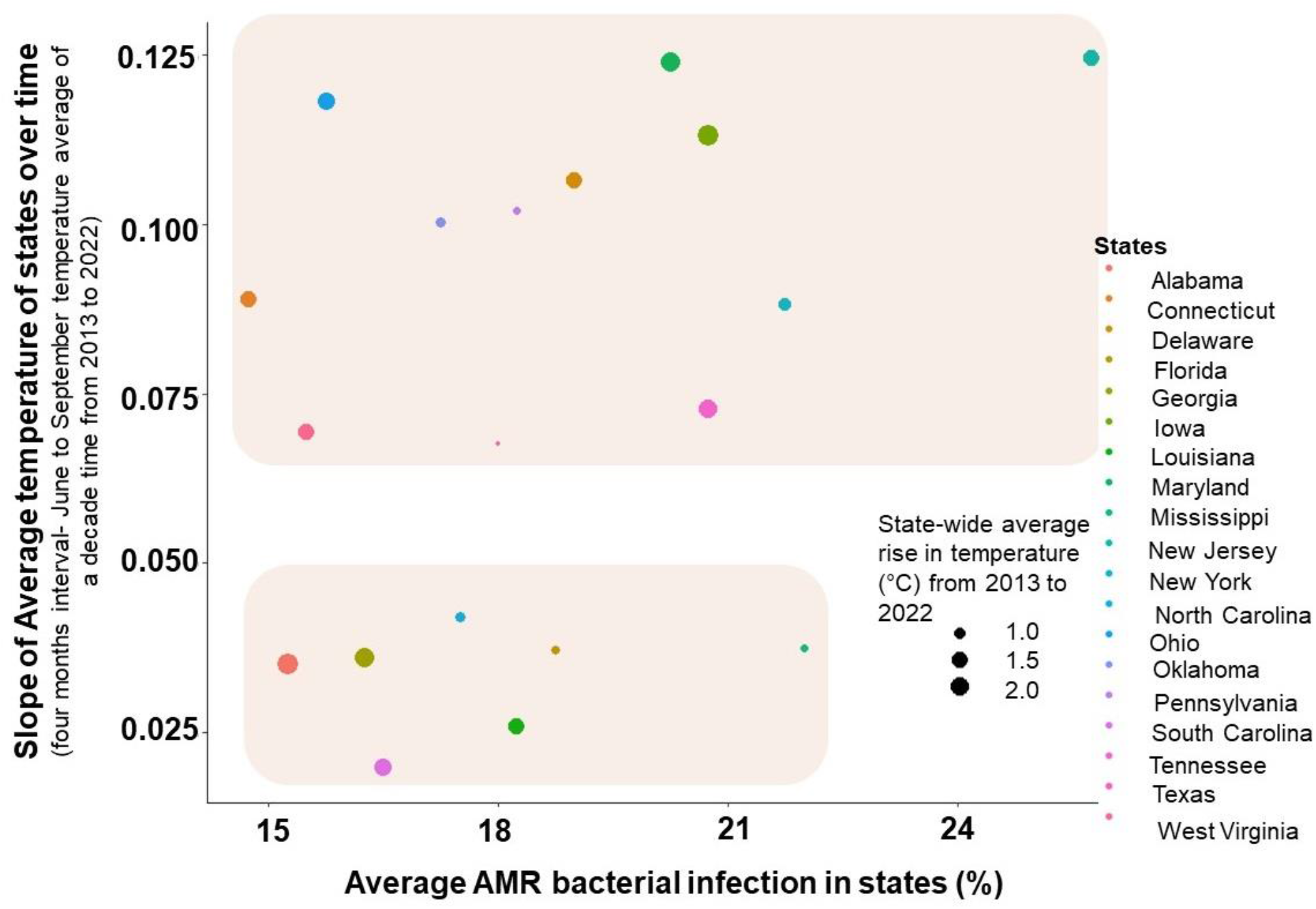
Generalized linear model computing the relationship between AMR bacterial infection and the rise in temperature in states in the United States, 2013-2022. The slope on the y-axis represents the slope of the linear model fitted for the average rise in temperature (°C) from 2013 to 2022 of each state. The state-wide average AMR bacterial infection is shown as a percentage. Regions shaded in the figure illustrate the clustering of states as a function of the increase in average temperature and AMR bacterial infections in states.

## 4. DISCUSSION

According to the study’s findings, there is a potential interaction between AMR, rising temperatures, and poverty. The higher rates of MRSA infections were strongly connected with a state’s percentage of Black or African American residents. Because of the higher risk of infection caused by insufficient access to healthcare because of poverty, AMR is more likely to appear in low-income, polluted states facing rising temperatures. Increased temperatures can also encourage the growth and spread of bacteria, particularly antibiotic-resistant ones. MRSA has been identified as a severe hazard to global health. Temperate climates, characterized by moderate temperatures and rainfall and favorable for bacterial development, are found in European, British, and Brazilian countries. Because of their milder climates, bacteria are potentially more vulnerable to acquiring antibiotic resistance in these nations’ higher AMR rates. The study also discovered that the high occurrence of MRSA infections was exacerbated by the state’s large black population and poor air quality in the United States. These results indicate a substantial relationship between AMR, poverty, and rising temperatures. Thus, it may be concluded that in the following ten years, AMR diseases like MRSA and VRE may be more prevalent in places with low income and high poverty. Furthermore, worsening environmental quality may result in a higher prevalence of AMR infections in states with poor air quality and rising temperatures.

Between 2013 and 2022, the GLM model anticipated the rate of AMR infection based on the number of bacterial isolates examined, the regions or locations of the states (geographic locations), and the average rise in temperature in the United States in the last 10 years. According to the model, the median value of an AMR bacterial infection may rise by 0.27% yearly. The model suggests that states with frequent shifts in temperature (remarkably increasing at a rapid rate of more than 1 degree per 10 years) might experience a surge in AMR infections in the future. However, the model also confirmed a high degree of variation in the data and that other factors, such as socioeconomic status and air quality, could have a vital impact on the development of AMR infections. The study also has several limitations-First, it was based on a cross-sectional study design, which cannot be used to establish causality. Second, the data used in the study was limited to factors that could contribute to the development of AMR infections, such as access to healthcare and hygiene practices.

Despite these limitations, the study provides valuable insights into the factors contributing to the rise of AMR bacterial infections in the United States. Most importantly, states like Alabama, Connecticut, Florida, Georgia, Louisiana, Maryland, New Jersey, New York, North and South Carolina, Ohio, Pennsylvania, and Tennessee are potentially more likely to experience an increase in MRSA and VRE bacterial infections due to a rise in temperature than other states. Overall, the study supported the hypothesis, stating the positive correlation between AMR emergence in pathogenic bacteria and changes in temperature. The current study provides insight into a less recognized but significant AMR and climate change nexus channel. It implies that additional research must be conducted to verify these findings and design effective prevention and management strategies.

## 5. CONCLUSION

In conclusion, this study found that socioeconomic factors, climatic changes, and racial differences affect the occurrence of AMR infections. States with the most significant risk of AMR infection were those with frequent temperature variations. As a result, climate change is projected to impact the dissemination of AMR considerably. The study also found a higher risk of AMR infection in states with higher poverty rates. The study’s findings demonstrate that comprehensive, multidisciplinary, and holistic approaches are required to address AMR and climate change issues.

## Data Availability

All data produced in the present study are available upon reasonable request to the authors

## SUPPLEMENTARY FIGURES

**Supplementary Figure 1.**
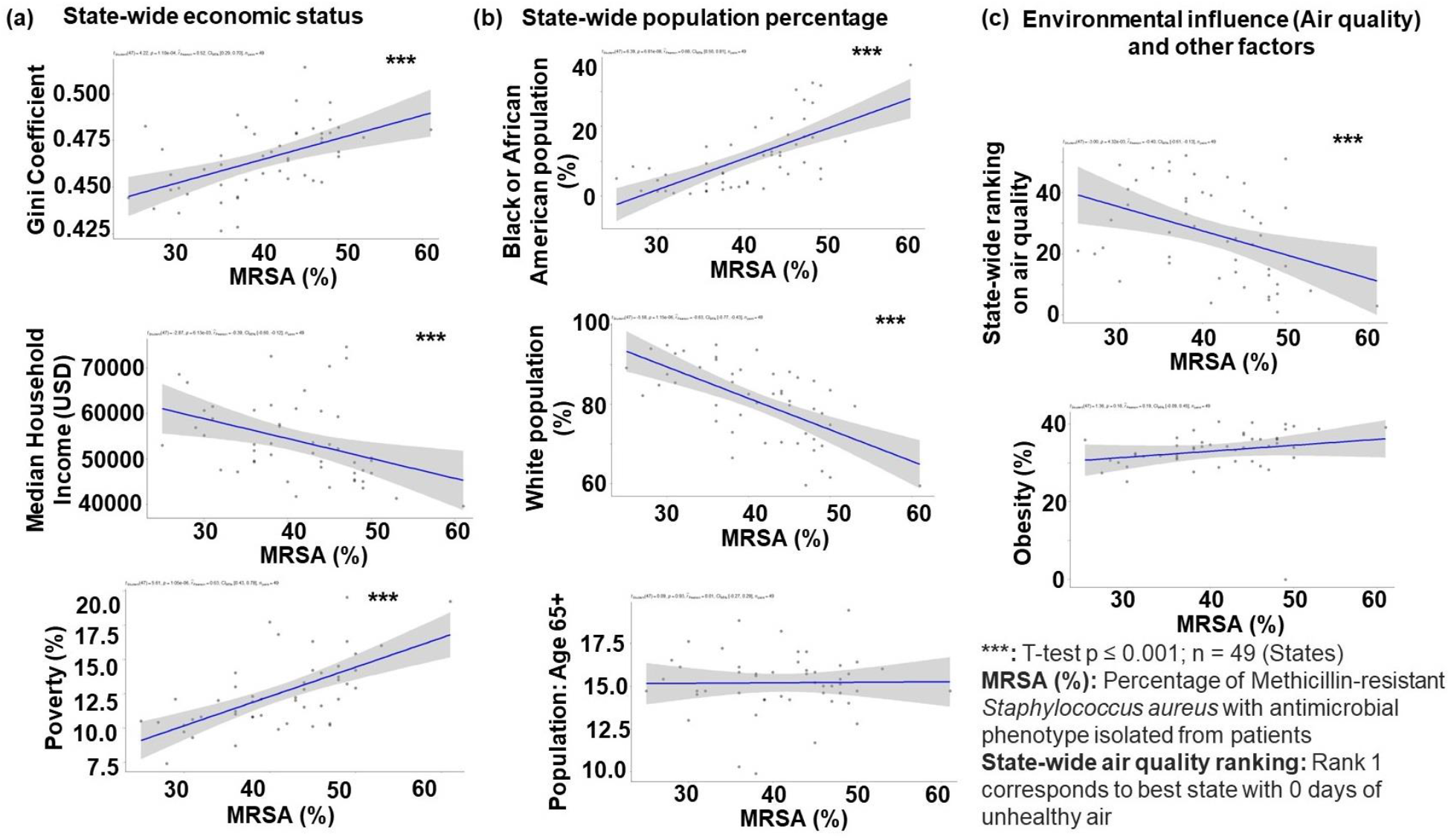
Linear regression of socioeconomic, population and environmental factors illustrating correlation with Methicillin-resistant *Staphylococcus aureus* (MRSA) infection in United States.

**Supplementary Figure 2.**
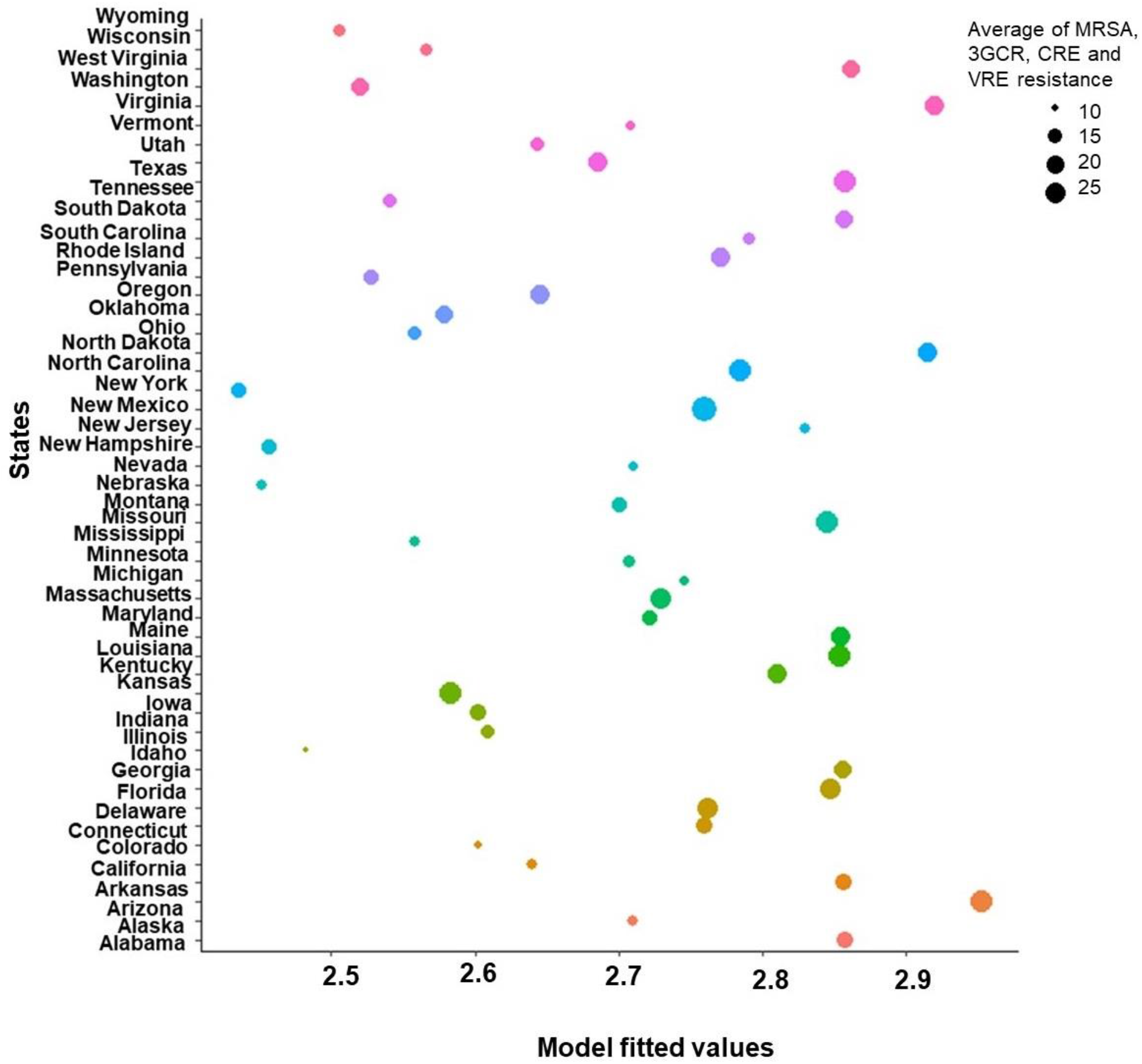
Generalized linear model fitted values of average AMR infection in United States

**Supplementary Figure 3.**
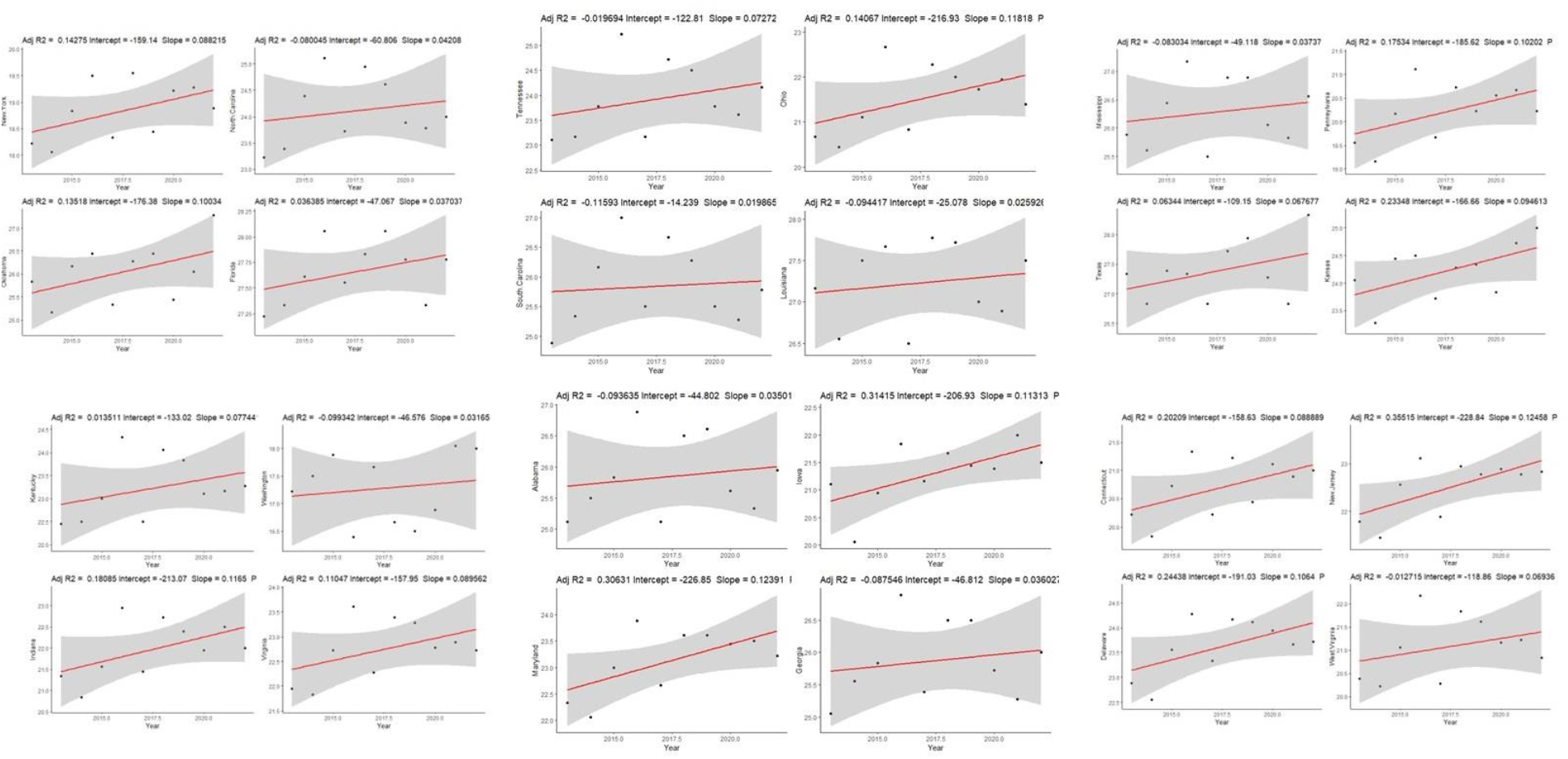
State-wide linear regression time series model illustration correlation of average temperature increase in last 10 years (2013 to 2022) in United States.

